# Conversational, Longitudinal, Ecological Assessment (CLEA): Exploring a new AI-driven method for qualitative data collection in a behavioural health context

**DOI:** 10.64898/2026.01.20.26344494

**Authors:** Samuel Downes, Thomas Krys, Kenton O’Hara, Max Western, Lauren Thompson, Amberly Brigden

## Abstract

In this paper, we present conversational longitudinal ecological assessment (CLEA), a novel conversational AI–enabled method for collecting ecologically valid, temporally sensitive qualitative health data via mobile instant messaging. We report findings from an exploratory deployment of an instantiation of CLEA within a 12-week community-based weight management programme, delivered by a charity partner in an area of deprivation. Using WhatsApp, we deployed our CLEA chat-agent to conduct twice-weekly conversational data collection sessions with participants, to elicit data about their experience of the programme and associated behaviour change. This was followed by in-person semi-structured interviews (N = 9) to examine user experiences and perceptions of interacting with the chat-agent. Participants reported that WhatsApp’s familiarity supported accessibility and sustained engagement, while the conversational format encouraged reflection directed towards the research focus. Responding to chat-agent prompts required cognitive effort, leading some participants to defer engagement until they had adequate time and mental space; however, this reflective demand was largely experienced as beneficial within the programme context. The AI’s quasi-human interactional qualities fostered a sense of support while reducing social judgement, enabling more candid disclosure. Together, these findings demonstrate initial feasibility and acceptability of CLEA for longitudinal qualitative data collection in an underserved population, and illustrate its capacity to elicit meaningful, contextually grounded insights consistently over time, that can be used in the formative stage of digital health intervention development. The study highlights both the opportunities and trade-offs of conversational AI for qualitative data collection, including design implications for health researchers looking to implement or extend the method. Finally, we position CLEA in relation to other longitudinal methods of health data elicitation.

**Author summary:** Developing effective interventions for health behaviours such as healthy eating and physical activity requires methods that can capture the complex, individual factors shaping people’s everyday experiences, including stress and motivation. Because such factors often fluctuate over time, longitudinal approaches are needed to understand how experiences and behaviours unfold in real-world contexts. For such methods to be effective, they must also be acceptable, engaging, and accessible—particularly for underserved or disadvantaged populations who are disproportionately affected by health-related conditions such as obesity. In this study, we introduce conversational longitudinal ecological assessment (CLEA), a digital health method that uses conversational AI technology to collect ecologically valid qualitative data over time through an accessible communication platform. We demonstrate the feasibility, acceptability, and utility of CLEA through a real-world deployment investigating an underserved group’s experience of a community-based weight management programme. To support other health researchers, we position CLEA in relation to existing longitudinal methods and highlight the key design considerations that shape engagement, data quality, and participant experience.

## 1 Introduction

Qualitative research is central to understanding complex health events, phenomena, and systems. Detailed insights into individual or group experiences can illuminate not just ‘what’ outcomes occur but also offer deeper understanding of ‘how’ and ‘why’ they occur [1]. Such experiences are subject to fluctuations over time, being affected by a complex interplay of personal, social and environmental factors that exert dynamic effects over a given period [2]. Eliciting the full depth and range of interacting attitudes, motivations, moods, emotions, behaviours and socioenvironmental context that shape people’s experiences over time is challenging. So to, is collecting data that is simultaneously insightful, relevant, ecologically valid and representative of an individuals’ authentic fluctuating experiences. This methodological balancing act must also account for the needs of respective participants. Researchers therefore face trade-offs in method selection, that varies according to research context, phenomenon of interest and resource availability.

### Methods for temporally sensitive, qualitative data collection

A common method for capturing temporal change is the retrospective interview—an in-depth conversation at a single time point that can produce rich, authentic narratives [3]. While skilled interviewers can build trust and elicit high-quality data through adaptive, dynamic techniques, data is reliant on participants’ recollections, afflicted by imperfect recall or social desirability [3]. Repeating interviews with the same participant over time can mitigate recall issues; though at significant resource cost. Additional methods can supplement interviews, such as collecting data from wearables, providing ecologically valid, longitudinal data to prompt recall [4]. Doing so, however, requires considerable setup costs, onboarding, and potential acceptability concerns around intensive tracking [5]. Beyond interviews, distinct intensive longitudinal methods exist for temporally sensitive research [6]. End-of-day diary studies (handwritten or digital) bound recall to a daily interval, capturing participants’ narrative summaries and interpretive appraisals of experiences unfolding across the day [6, 7, 8]. Considering data provision is self-led, it can vary in relevance and clarity, increasing analytic demands for researchers [9].

Another longitudinal research method, ecological momentary assessment (EMA), involves prompting participants more frequently about momentary experiences, to elucidate fast, transient, and context-sensitive phenomena [10]. To enable this, many designs rely on constrained response formats (e.g., short-answer items or quantitative scales); with the trade-off of limiting richer disclosure [11, 12]. EMA also typically requires considerable onboarding, and close compliance management due to the burden imposed on participants by the prompt frequency [10]. In studies using EMA for behavioural health research, compliance has been demonstrated to decline linearly over study duration [13]. In addition to extending the acceptability and engagement of longitudinal methods, methodological value could lie in methods that combine the interpretative richness of end-of-day diaries, with the directive, topical focus characteristic of EMA designs. Such methods could be appropriate for research areas such as behavioural health research, where individually complex determinant factors fluctuate over time and are strongly shaped by personal context; and where EMA designs may miss important contextual nuance or interpretation [14, 15, 16].

### Conversational agents (CAs) for qualitative data collection

There has been growing interest in recent years in the application of artificial intelligence (AI) to qualitative research. While much prior work has focused on using AI to support qualitative data analysis [17, 18 19], comparatively little attention has been paid to how it might be used in data collection. This is surprising considering the potential of conversational AI to collect data in a directive, engaging manner that makes space for interpretive richness. Initial studies have demonstrated the feasibility of AI-powered conversational agents (CAs) for conducting qualitative interviews or inquiry, most often in single encounter or non-naturalistic laboratory settings [20, 21, 22, 23]. To date, however, there has been limited investigation into the deployment of CAs for longitudinal, in-the-wild qualitative data collection, or their methodological implications relative to EMA, diaries, or interviews. A brief overview is provided below of CA technologies and existing studies exploring their potential in qualitative data collection.

### Evolution of conversational agents (CAs)

CA technologies have co-evolved with developments in AI over the last century. Early systems used hand-crafted rules to emulate a sense of conversation, albeit highly constrained [24]. An example of such a chatbot deployed for qualitative data collection is Tallyn et al.’s “Ethnobot” [25], used to gather experiential, momentary qualitative data during a public event. While users of this early system frequently anthropomorphised it, many were disappointed by its repetitive behaviour or frustrated when it did not meet their conversational expectations—a phenomenon described as the “gulf in user expectations” [26]. More advanced CA technology combines machine-learning capabilities alongside older rules-based technology, in hybrid systems [24]. Xiao et al. [20] compared performance of a hybrid CA to traditional fixed-question survey, with participants (video-gamers) providing feedback on video-game trailers. They gathered more informative data from the CA enabled tool, and participants expressed greater positive sentiment towards the CA. While video-gamers are more likely technophiles, this work nevertheless highlights how hybrid CA systems can be effective for qualitative data collection within pre-specified topics, potentially outperforming traditional fixed-question surveys.

More recently, with the emergence of large language models (LLMs), CA systems are now capable of more dynamic conversation, sustaining coherent, dialogue with greater generalisability to different topics and use-contexts [27]. Several studies have investigated their use for qualitative data collection. A large-scale study (n = 399) comparing CA technologies for qualitative data elicitation found that despite simpler development requirements, a lightweight LLM-based CA elicited similarly informative data and was more favourable to users than a sophisticated hybrid CA [28]. Geiecke and Jaraval [25] conducted a rigorous examination of a lightweight LLM-based CA by recruiting sociologists to score transcripts of AI-led interviews, reporting data obtained as on-par to an “average human expert” interviewer. They also found participants expressed preference for AI-led interviews over human interviews, as the AI was non-judgemental.

Advances in the conversational ability of CAs have demonstrated the possibility of conducting engaging, single-session conversational data collection remotely at scale [20, 22, 23, 25]. The work of Xiao [20], Cuevas [28] and Geiecke and Jaraval [21] highlights how these newer forms of CA technology may narrow the gulf in user expectations that limit richer disclosure in earlier generation CAs, like Tallyn’s ‘Ethnobot’. The previously detailed works demonstrate that participants provide more informative, detailed responses relative to static, traditional survey approaches. Whether this extends longitudinally, to methods such as EMA or diaries in health contexts is yet to be established.

To address this research gap, we present an exploratory deployment of a new method for conversational, longitudinal, ecological assessment (CLEA), that utilises conversational AI and mobile instant messaging. We position CLEA as a high-level ILR longitudinal research method that, through specific design choices, can be adapted to conduct longitudinal research with varying qualities of EMA or diary method depending on requirements of the respective study context. In this work, we explore the use of CLEA for qualitative data collection in a behavioural health context. The presented instantiation of CLEA employs LLM-based conversational AI and WhatsApp to gather twice-weekly qualitative data over a 12-week weight management health intervention programme (henceforth referred to as ‘the programme’) in an area of deprivation.

Our research aims in this work were to establish A) feasibility of the tool B) acceptability of the tool to users, and C) the utility of the data captured over this period. We adopt a pragmatic approach for guiding methodological decisions that embraces both a constructivist consideration of knowledge production and an interpretative lens for qualitative analysis. Through our findings, we evidence the potential value of this technology-mediated method as well as its constraints. Drawing on this analysis, we contribute early insights into the design of next-generation tools for conversational longitudinal ecological research.

## 2 Methods

### 2.1 Study Design

A mixed-methods study, analysing chat log data (from participant engagements with CLEA) alongside qualitative interview data to explore the acceptability, feasibility and utility of the novel CLEA system.

### 2.2 Setting and context of CLEA deployment

This study was conducted in collaboration with the Robins Foundation - a local charity associated with Bristol City Football Club, providing health and wellbeing support to underserved communities in Bristol, UK. The Foundation deliver a 12-week behavioural health programme, targeting weight loss, in an area of deprivation. Within this programme, we deployed an early version of CLEA, to gather data regarding (1) participants’ immediate feelings after attending a health session to understand their experience; and (2) the barriers/facilitators participants experience in applying session knowledge and skills in their daily life. Features of this instantiation of CLEA were tailored according to collaborator advice, providing alignment to the needs of programme participants. Through this, we aimed to shed light on the acceptability (in terms of participant experiences, perceptions and attitudes), feasibility (terms of instrument fidelity and data completeness), and utility (how useful the data is over time) of conversational, longitudinal, ecological assessment in a behavioural health context.

### 2.3 Participants and recruitment

Individuals were eligible to participate if they were: (1) at least 18 years old, (2) English-speaking, (3) had access to a smartphone with WhatsApp, (4) were in stable housing, (5) no critical health conditions and (6) engaged in the programme. Baseline socio-demographic information was collected, and participants received onboarding instructions for the CLEA system. Sampling was opportunistic via the programme, with potential participants provided with study information at week 1, invited to provide informed consent from week 2, and subsequently onboarded from week 3. Individuals could join the study up to halfway into the programme.

### 2.4 CLEA system

Through guidance from our charity partners, we prioritised accessibility in the design of this instance of CLEA. With that in mind, we selected WhatsApp as the interface for participant interaction with the CLEA chat-agent. WhatsApp is a commonly used messaging application that is easy-to-use, cheap, and familiar to the majority of iPhone and Android smartphone devices.

Being available over WiFi networks, WhatsApp enabled cost effective use for those without cellular data plan. Furthermore, onboarding was straightforward, identifying users via their telephone number rather than requiring a password-protected account. Partners endorsed this as an inclusive choice for user interface; they themselves use WhatsApp for group communications, finding it to be highly accessible to those with lower digital literacy. The WhatsApp interface can be seen in Fig 1.

**Fig 1.**
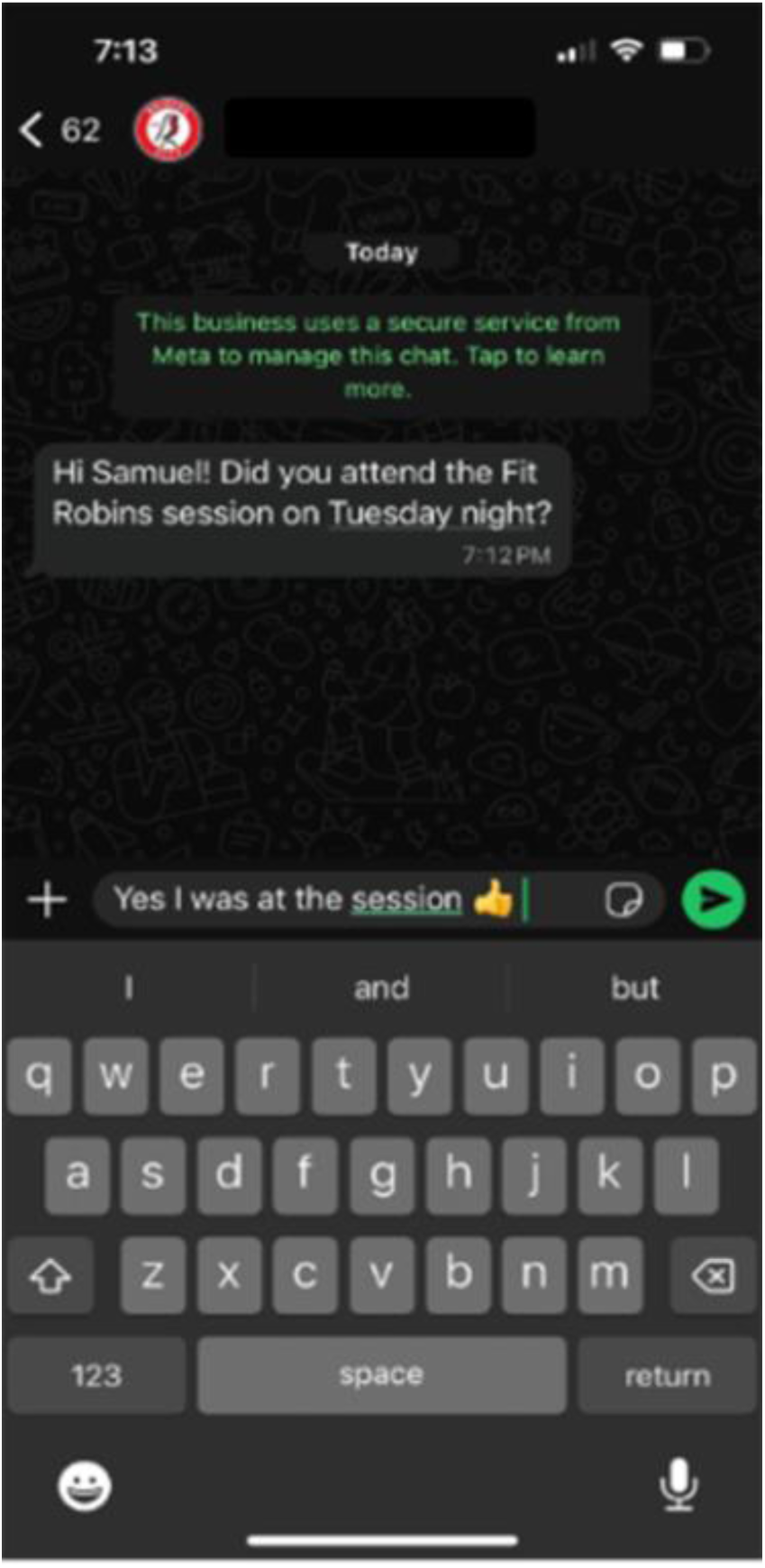
Sample visual of user interface in responding to a single data collection prompt by the CLEA system in WhatsApp

The WhatsApp front-end enables participants to send and receive messages to our chat-agent, via our custom web application, which forwarded messages to and from our selected LLM, OpenAI’s GPT4-Turbo. Chat-logs from these messaging sessions would be periodically downloaded via an encrypted connection, to a password protected Excel spreadsheet hosted on the University’s encrypted Microsoft SharePoint site. In terms of piloting the acceptability of the AI, partners appreciated its generally supportive tone, feeling it would be appropriate for programme participants.

The choice of LLM ’under the hood’ is fundamental. At the time of development (2023), OpenAI’s GPT4-Turbo model was preferred over alternative models that were still nascent and less performant. The GPT4-Turbo model tends towards a positive, affiliative tone and leans towards ‘safe’ interactional practices - avoiding overly sensitive or emotional topics [29, 30]. This tendency towards ‘safe’ interactions was desirable in this instance, considering typical in-person ‘distress protocols’ such as reading non-verbal cues are not applicable here [31].

Our application contained specific prompt-based instructions to guide the AI in managing conversation flows with participants. Figures 2a-c detail the core operating instructions and research targets for the question-answer sessions. The chat-agent was instructed to act as a reflective interviewer, positioning it to elicit personal reflections. The prompt instructions prioritised understanding the participants’ feelings, experiences, and behaviours. This, in effect, guided the content and frames of dialogue. Based on partner advice to manage participant burden and prioritise accessibility, the chat-agent was instructed to use simple language, ask no more than five questions and switch sub-topic after a maximum of three questions within a session.

**Fig 2.**
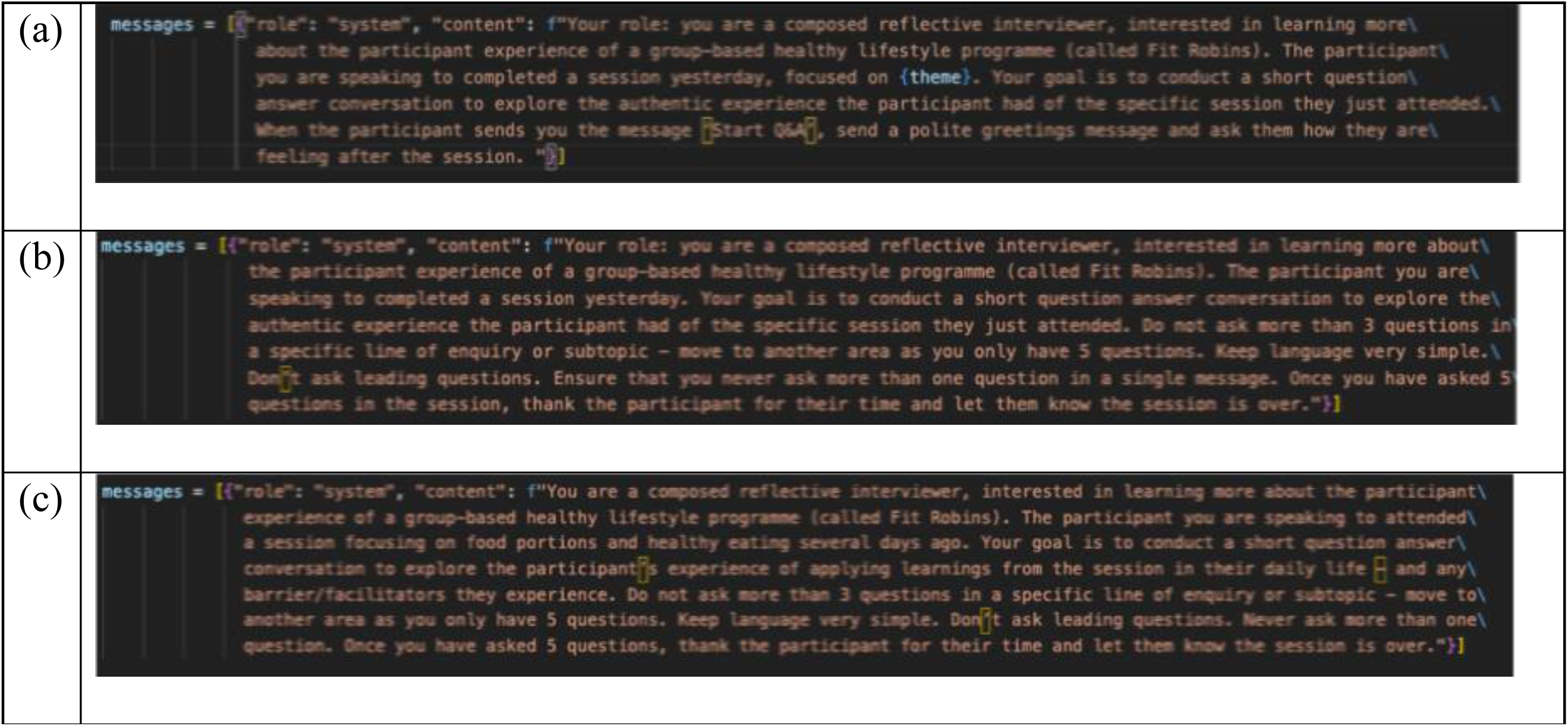
**(a) Post-session reflection data prompt – e.g., on the Tuesday after attending the programme Monday night – to explore participant experiences through their feelings about the session, (b) Continuing post-session reflection data prompt - to explore further participant experience of the session, (c) Follow-up weekend prompt – to explore participant enablers and barriers of behaviour change in the days following a programme session.**

Additionally, the system was instructed to ask one question at a time and not lead participants. Finally, chat-agent-generated messages were capped at a maximum length of approximately 75 words. Throughout development, we sought to balance rich data collection with the burden placed on participants. Other choices and trade-offs might be considered more appropriate in different contexts of CLEA usage.

### 2.5 Data collection and analysis

To contextualize our data collection and analysis we consider the following research aims and objectives:

- **Aim A:** To determine the feasibility of CLEA for real-world longitudinal qualitative data collection, in terms of instrument fidelity and data completeness

■ **Objective A1**: To assess chat-agent question clarity and relevance to the instructed research target (fidelity)
■ **Objective A2**: To assess participant longitudinal engagement with CLEA, in terms of number of data collection sessions responded to, sessions completed, frequency of deferred responses and response length (within and across participants).
- **Aim B:** To evaluate the acceptability and perceived usability of CLEA

■ **Objective B1**: Explore participant experiences, perceptions and attitudes to chat-based interactions with CLEA
- **Aim C:** To determine the quality and utility of longitudinal qualitative data gathered using CLEA

■ **Objective C1**: To understand CLEA’s ability to collect new and insightful data over time per participant
■ **Objective C2**: To assess clarity, relevance, and approximate reflective depth in participant responses to CLEA’s questions
■ **Objective C3**: To explore participant experiences and perceptions to the provision of meaningful data over time, and the alignment to observed behaviours in chat-logs

#### Chat-log data analysis: Exploring the feasibility and utility of CLEA (aims A and C)

Full chat logs of the chat-agent/participant interactions were extracted from our database (see 2.4) for all participants over the study period. The active study period took place from week 3, through week 12 of the programme, yielding 18 possible data collection sessions. with the aggregated dataset totalling 555 chat-agent messages and 572 participant messages (occasionally participants sent multiple responses to a single chat-agent message), across 18 possible data collection sessions. Only 18 data collection sessions occurred over the study period, due to the first three weeks being used to recruit participants, and a single week where data collection was attempted once, as a programme session was not held that week.

Quantitative analysis of this log data was undertaken, providing descriptive statistics on the number of completed chat-agent data collection sessions, partially completed sessions, uncompleted sessions, as well as analysing length of responses; both at the aggregate and individual participant level. These insights were used to answer research objective A2.

Inductive content analysis of chat-logs was conducted per participant to identify prevalence of new codes over time relative to the research target (see 2.2). TK and SD each coded a single participant’s chat-log data, meeting to discuss codes and gain interpretative consensus, per Hsieh and Shannon [32]. SD then completed the coding of the logs for the remaining participants. Outputs were used to understand the feasibility of collecting useful longitudinal data with CLEA (C1).

Relevance and clarity scoring of 1) chat-agent messages and 2) participant responses to chat-agent questions was performed. These metrics are adapted from Grice’s Maxims for information quality, operationalised in similar analyses [20, 28, 33]. Scoring of chat-agent messages is novel, not being pursued in similar analyses by Xiao [20] or Cuevas [28]. AB, TK and SD each scored relevance and clarity for two full WhatsApp transcripts, before comparing results and resolving minor discrepancies within a single meeting. For subsequent transcripts, TK completed scoring for chat-agent messages, and SD for participant responses. Details regarding scoring practice are available in Table 1. Scoring was used to answer research objectives A1 and C2.

**Table 1:** Metrics for scoring clarity and relevance of messages.

| How codes were derived | Code definition | Scoring |
| --- | --- | --- |
| Top down: adapted Gricean Maxims [33] | **Clarity (adapted from Manner maxim)** of chat-agent messages and participant responses -considering succinctness, ambiguity, repetition, coherence | 0 (unclear), 0.5 (some ambiguity, repetition, superfluity, or incoherence present), 1 (fully clear) |
|  | **Relevance (adapted from Relation maxim)** of chat-agent messages to the research question, and of participant | 0 (irrelevant), 0.5, (contains both relevant and irrelevant elements), 1 (fully relevant) |
|  | responses to chat-agent questions. |  |
| Reflective depth | Emotional, and or cognitive reflection demonstrated by a participant in response to a chat-agent question, focusing on the extent to which participants connected thoughts and emotions to experiences in their responses. | 0 (Descriptive reporting of events or states with no explicit emotional response or cognitive appraisal), 0.5 (Inclusion of either an emotional response or a cognitive appraisal, without integration of both), 1 (Integration of emotional response *and* cognitive appraisal in relation to the reported experience) |

Through the outlined analyses of logs, we noticed variations in the apparent depth of answers provided by participants. Therefore, we decided to include this in our analysis. This was operationalised through an approximate heuristic of ‘reflective depth’, constituting the level of cognitive and or emotional depth perceptible in participant responses. AB and SD each scored two full transcripts, following the same process as (3) above; SD subsequently completed scoring for remaining transcripts. Scoring contributed to objective C2.

#### Interviews: Exploring the acceptability, usability and utility of CLEA (aims B and C)

At the end of this 12-week period, participants were invited to a one-off, semi-structured, in-person qualitative interview (average duration of 40 minutes) conducted by SD. All participants who completed the 12 weeks consented to participate in this interview. Interviews explored participant impressions of using the chat-agent, views on WhatsApp, attitudes towards the AI, experiences of interaction, preferences, motivations to engage or disengage, and perceived value of the system (see Appendix A for the topic guide). Qualitative interviews were audio-recorded and transcribed verbatim.

Three members of the research team (SD, AB, TK) conducted a thematic analysis [34] using NVivo 20. First, each researcher independently coded a subset of four transcripts, regrouping to reach consensus on an initial inductive codebook. SD and TK then coded the remaining transcripts, meeting to discuss and resolve discrepancies; refining codes as needed. SD and AB iteratively clustered codes into themes and sub-themes, with feedback from KOH. As the research questions were: 1) specific (experiences of using chat-agent), 2) the group were relatively homogenous (from a similar socio-economic background and enrolled in a weight- related health programme), and 3) the interviews were rich; the sample was found to have sufficient information power [35] to answer objectives B1 and C3.

### 2.6 Ethics and data protection

The technology system was approved under a University of Bristol Data Protection Impact assessment. This included guardrails, such as manual daily monitoring of conversational logs in the interests of participant safety. The study received University ethics approval (ref: 14542-17493). Ethical issues included giving particular attention to the accessibility of the participant information sheet and consent forms, given the potential for lower literacy levels in the participant group. During the study, conversation data was stored in a private encrypted database (see 2.4). After the study, chat log and qualitative transcripts were stored in a secure location within the university’s digital environment accessible only to the core research team. Any identifiable data was redacted (pseudonymisation). Participants were informed that data would be held for up to 10 years, and they have the right to request the deletion of their raw data up until the point of analysis.

## 3 Results

### 3.1 Participant characteristics

Nine participants joined the Robins 12-week programme. From this cohort, we recruited all nine participants with a mean (SD) age of 46 (16.5). Five participants joined on week three of the programme, three on week four, and one on week five. Seven identified as female, and the sample was predominantly White British (n=7). Three participants fell into the lowest decile of average gross household UK income, three in the second-lowest decile, two in the third, and one in the fourth. Two participants had secondary school as their highest level of education; six had completed college, and one had attained a university degree. All participants had a smartphone and familiarity with WhatsApp. All nine participants were interviewed at the end of the study.

### 3.2 Relevance, clarity of chat-agent questions (Objectives A1)

Across 555 questions, relevance and clarity were high (relevance 0.99/1; clarity 0.99/1). This highlights the chat-agent’s ability to formulate clear questions focused on the target research phenomenon set out in its instructions (see 2.4), indicating good fidelity.

### 3.3 Exploring participant engagement to establish feasibility for longitudinal data collection

#### 3.3.1 Overall engagement statistics (Objective A2)

Fig. 3 demonstrates the longitudinal feasibility of our chat-agent for data collection. Across all participants, data was collected for 80% of scheduled sessions. 102 data collection sessions (green) were completed in entirety, 13 (yellow) were partially completed, and 29 (light grey) were not answered. 18 sessions (dark grey) lacked data due to participants entering the study at different times, along with a system outage resulting in a lack of data collection for message block 6. When participants engaged in a data collection session with the chat-agent, 101/102 sessions were completed within 3-7 minutes.

**Fig 3.**
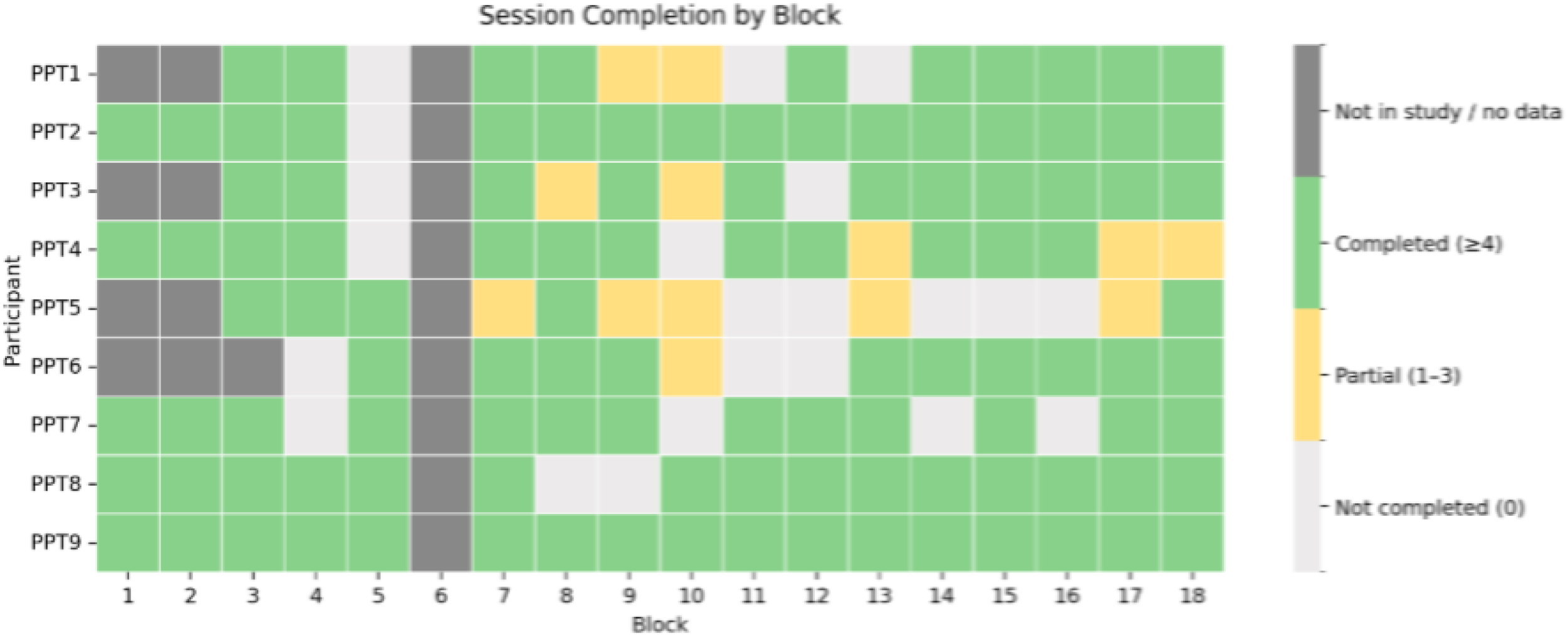
Longitudinal participant engagement over study period, showing data prompts completed in full, partial completions and non-completions; message blocks 1 & 2 take place on week 3 of health programme.

An important consideration for understanding the temporal sensitivity of the method is understanding the extent of delayed responses by participants. Fig 4 illustrates the deferral patterns of participants, with data provision deferred 12 hours or more at least once for all but one participant. In aggregate, data provision was deferred on 33% of occasions. This highlights the flexibility of the method, as well as the tension between momentary validity and participant acceptability.

**Fig 4.**
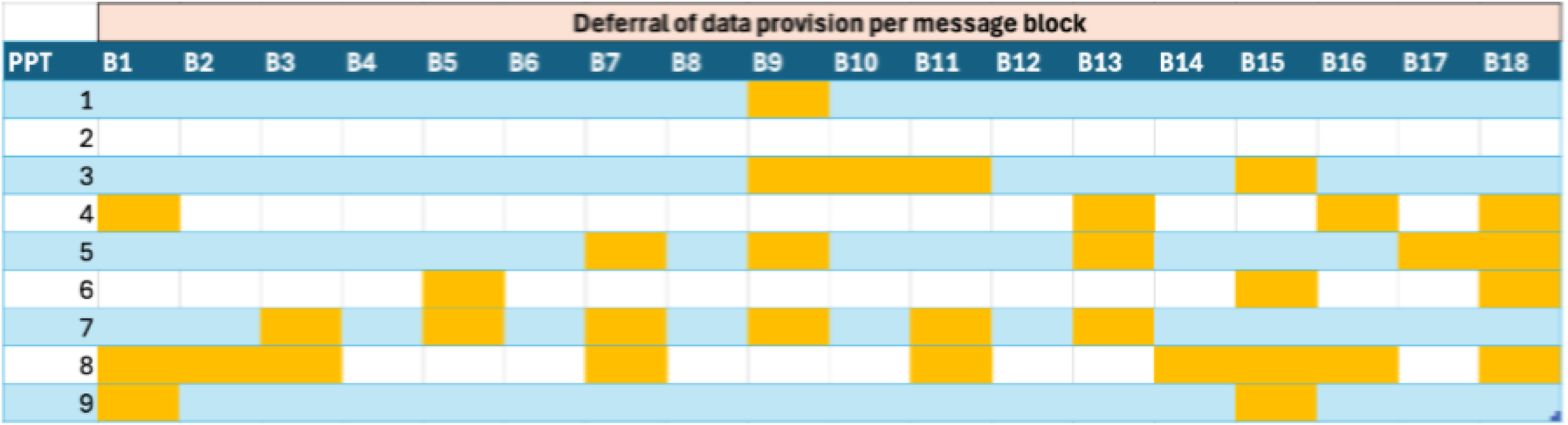
Prevalence of message blocks (B1, B2 etc) that were deferred per participant. Amber cells indicate deferral of 12 hours or greater from the point at which a prompt for data collection was sent to when a response was received.

Across the nine participants, a total of 572 responses were received, displayed by response length per participant in Fig 5. The mean length approximated to 7-9 words. The maximum length of responses per participant varied substantially (approx. 20-60 words) and averaged around 30-40 words. This indicates responses were typically brief, however the displayed variance shows this could be shorter (one word) or, on particular occasions, substantially longer.

**Fig 5.**
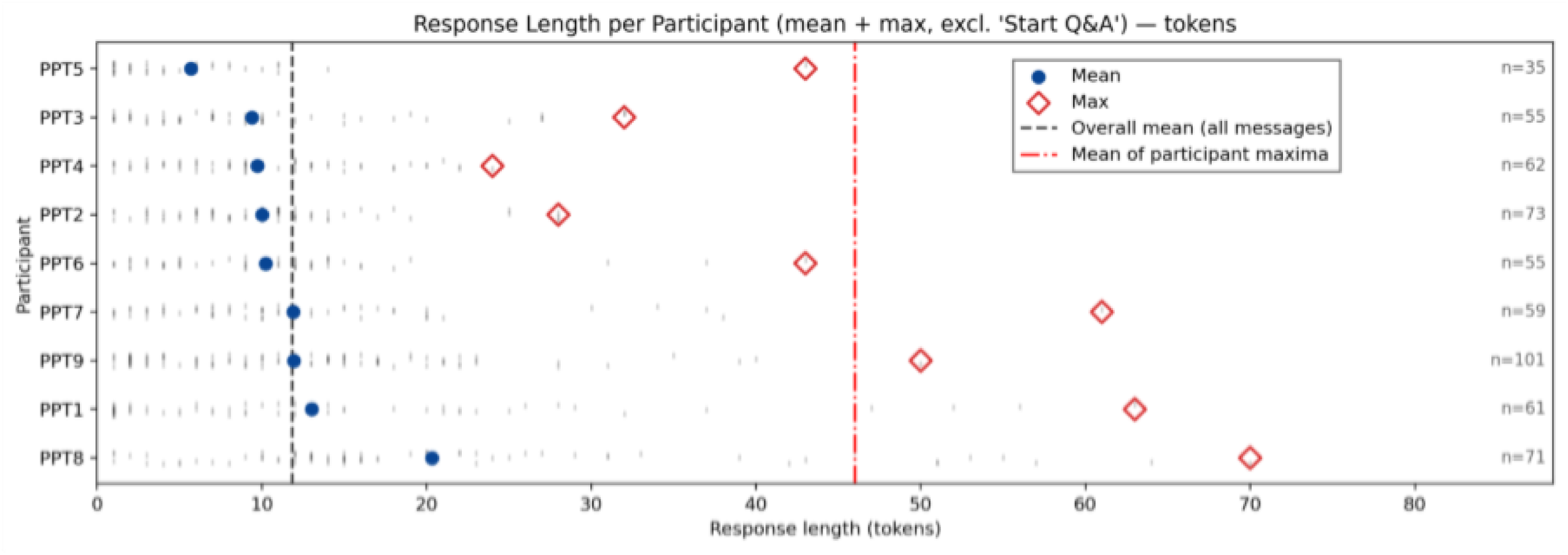
Average participant response length (tokens) across all received responses, displaying 1) per participant mean, 2) per participant maximum response length, 3) overall mean for all participants across all messages, and 4) pooled mean of maximum response lengths.

#### 3.3.2 Users experiences and perceptions of engaging with CLEA (Objective B1)

##### Theme 1: The WhatsApp Interface: Frictionless, Easy and Accessible

No participants reported experiencing issues accessing the chat-agent through WhatsApp. All participants already had WhatsApp installed and none reported technical issues answering the twice weekly Q&A sessions. This is supported by the session completion data (see Fig 3) with all but one participant successfully responding to their first Q&A session (PPT6, Block 4).

Participants were positive about the ease and accessibility of the WhatsApp interface. Indeed, one participant who self-identified as ‘t*ech-averse*’ described their experience of responding within WhatsApp as ‘q*uick, easy and painless’ (PPT 8),* acknowledging WhatsApp is ‘*second nature’* for many people, speaking to it being broadly familiar and accessible. Participants appreciated how the interaction with WhatsApp was easy, utilising the simple click through from a notification to providing a text response - ‘*It’s something that comes directly through to the phone and you don’t have to do anything else other than just text or reply’* (PPT 6).

##### Theme 2: Value of engaging with system compensates for cognitive effort required

While the interface itself posed few barriers, several participants noted the cognitive effort required to thoughtfully engage in answering reflective questions. They expressed a desire to ‘*focus’ (PPT7)* to answer the question carefully, suggesting a requirement for a suitable psychological state and physical/social setting. As such, participants often self-selected appropriate times to answer - ‘*Sometimes I’m not in the right place [headspace] to answer*,” and ‘*I don’t respond to it until I know I’ve got time.’;* often deferring answering ‘*until the evening when I’m sitting down after supper and I can actually think about it carefully’ (PPT9), ‘ignore it and wait until I get home’ (PPT7)*. This reported deferral is supported by the chat-log data, where some participants’ Q&A sessions were unanswered at the point of initial prompting, but were answered later that day or the next (see Fig 4). This phenomenon therefore had connotations for the temporality of responses. However, some noted this flexibility of data provision could lead to forgetting to complete it - ‘*a lot of the time it arrived when I was in the middle of something really busy, so my initial response was kind of that’s going to have to wait, and then sometimes I would forget and it would be a couple of days that it would be waiting, rather than the hour or so that it needed to’ (PPT6)* - resulting in missing or delayed data.

While perceived cognitive challenge could be an acute barrier when receiving prompts, participants generally perceived engaging in the Q&A sessions to be beneficial. Participants described prompts encouraging active recall and reflection on session material, encouraging them to ‘*go back and think’* and ‘*remind’ (PPTs 2/8/9)* themselves of the programme’s content. Participants reported an interventional effect of this cognitive reflection in making the programme information more salient and stimulating consolidation of learning (‘*It makes you then think about what you’ve learned’ (PPT1), ‘It’s a way of reminding yourself what you actually talked about … and kind of makes you think how you are going to apply that’ (PPT8)).* Participants described further interventional effects, whereby engaging in a Q&A session prompted self-monitoring of their health behaviours, sometimes resulting in action towards behaviour change- ‘*It does make you kind of reflect on programme content’ (PPT8), ‘it’s a reminder about what you know so the next day to you can just stay focused, just keep going.*.’ (*PPT2*). In the context of the programme, the investment of cognitive effort therefore provided value to participants, promoting sustained engagement longitudinally.

##### Theme 3: Sense of social support and rapport fostered by the chat-agent

Participants further noted a sense of psycho-social support from interacting with the chat-agent, describing this support as another factor in their sustained engagement. Participants expressed sentiments such as ‘*It gives you a sense of support…’ (PPT2)* and ‘*you’ve got somebody to talk to’* and described forming an ‘*important*’ *(PPT9)* connection with it. Participants cited the positive, encouraging, and reinforcing manner as important in fostering a sense of support and positive feeling — ‘*It made me happy because it was giving me positive feedback’ (PPT2)*.

Furthermore, this affiliative capacity encouraged honest disclosure - *‘the connection I think is also important, for fostering honesty*.’ *(PPT2)*.

The sense of support and emotional connection was related to participants feeling that the chat-agent listened to and understood their responses (‘*It’s almost like it’s reading it back to you and you think ‘yeah, that is right’…*’, ‘*It listens and pays attention*’, ‘*It knows m*e’ *(PPT9); ‘So it’s*

*like someone listening’ (PPT1))*. Furthermore, one participant described a sense of validation - ‘*It feels a bit more validating’ (PPT9) -* when it reflected back their thoughts and feelings. In addition, most participants praised the system’s manner and immediacy of response, suggesting it felt like a ‘r*eal-time’ (PPT9), ‘engaging experience’* (*PPT8*), and that ‘*with the chatbot, it just sort of flows*’ (*PPT4*). However, some participants did acknowledge occasional moments where the chat-agent seemed more robotic, repetitive, or lacking memory of what participants had said in prior sessions, which could undermine their engagement.

While participants had varied attitudes towards conversing with *‘just a bot’ (PPT2)*, this did not prevent them from engaging in rich conversation: ‘*It doesn’t matter if it is you [human researcher] or a chatbot because the responses are picking up on the text that I’m writing… it is just as good’ (PPT9)*. The variation in responses suggests that the extent to which participants engage in natural conversation with a chat-agent may be influenced by pre-existing attitudes or perceptions towards such technology.

### 3.4 Exploring Longitudinal Utility (Objectives C1-3)

#### 3.4.1 Longitudinal utility of data collected through the chat-agent (Objective C1)

The heatmap in Fig 6 shows that the presence of new codes was consistent over the 12-weeks, indicating the chat-agent was able to yield new and relevant insights per participant over the study period. This provides evidence that CLEA can gather useful insights into phenomena of interest as they fluctuate over time. Examples of codes are included in supplementary material (S1).

**Fig 6.**
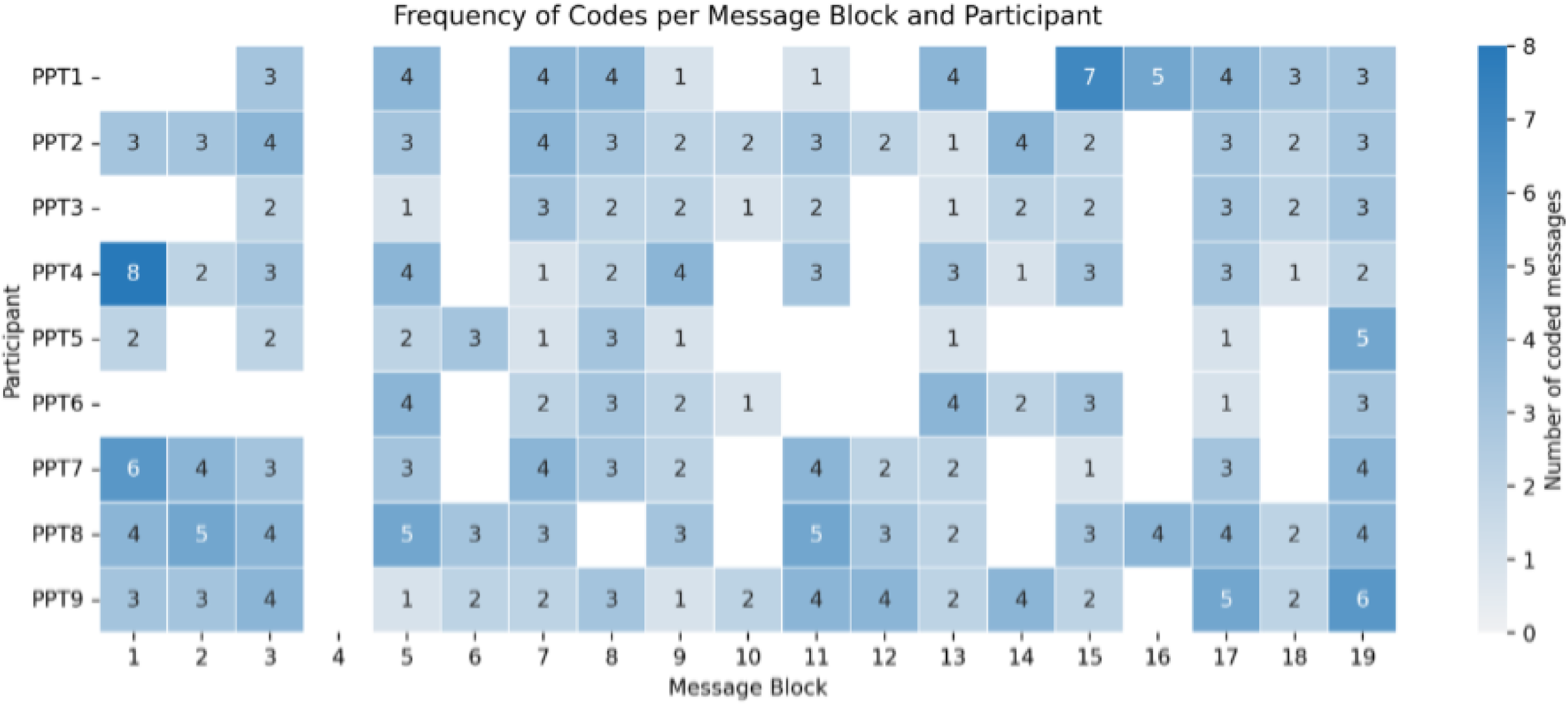
Heatmap of new, relevant thematic codes plotted over study duration per participant per message block

#### 3.4.2 Relevance, clarity and reflective depth of participant data samples (Objective C2)

Relevance of responses provided by participants to the chat-agent was 0.95/1, demonstrating that participants understood the meaning of questions asked, and could respond with relevant information. Clarity of responses was 0.89/1, meaning the majority of data provided by participants was coherent and succinct. Our approximate heuristic of reflective depth averaged 0.46/1, indicating participants’ typical single responses were sparing in cognitive or emotional depth. Additionally, content was found to typically contain short and specific descriptions of recent behaviours, emotions and events.

#### 3.4.3 Participant experiences and perceptions relating to data quality over time (Objective C3)

##### Theme 4: The tension of neutrality and rapport-building

As reported in theme 3, across interactions, the chat-agent was perceived to utilise positive language, encouraging statements and positive reinforcement. Sometimes these were general statements of encouragement or positive reinforcement for sharing information, akin to general rapport-building techniques used in qualitative interviewing (‘*that’s insightful’ [AI], ‘Interesting!’ [AI], ‘Thank you for sharing that personal experience.’ [AI], ‘I appreciate your openness.’ [AI]*). More commonly, the encouraging statements were non-neutral, endorsing the opinions, behaviours or feelings expressed by the participant, ‘*It was giving me positive feedback*’ (*PPT2*). This non-neutral feedback appeared to have an interventional quality, tying into the sense of social support outlined in theme 3.

Statements of encouragement rarely led to greater expansion or depth of response from participants. While unconditional positivity was common (and typical of the underlying GPT-4 turbo model), we found no instances of the chat-agent using techniques to challenge responses, such as techniques to explore apparent contradictions, with connotations for data quality. Indeed, we found participants tended towards positive sentiments about the programme and their health behaviours. It is possible that the participant’s positive sentiments were reinforced by the chat-agent and/or its positive manner, through setting the tone for interactions. While the positive encouragement may have created an issue with shaping the data, some users reported this characteristic to promote a sense of non-judgemental, enabling richer disclosure - ‘*I think it potentially means you can be…a little bit more honest…because you’re not thinking about being judged when it’s an AI*’ (*PPT2*).

##### Theme 5: Interactional practices of the chat-agent affecting data quality

Participants drew direct comparisons between the chat-agent’s ability to engage in an open conversational style and more traditional qualitative surveys: ‘*We’ve all filled out those surveys where your answer doesn’t fit any of the boxes… at least your questions [CLEA] require a personal answer’*. The perceived personal and conversational nature was reported by some participants to lead to more ‘*in-depth’* and ‘*detailed’* responses: ‘*It makes you answer more in depth because you feel like you’re talking to a person…’.* Some also felt this allowed greater scope for sharing authentic personal experiences: ‘*It definitely makes you think that what you’re saying is important, whereas with a generic questionnaire, I think ‘oh, not another one.’ (PPT9)*.

Many participants explicitly cited the lack of a real human as a reason for providing more candid responses. *“I suppose [with a human] I’d be a bit more careful about what I ought to say, but with the chatbot, it just sort of flows…”.* Another participant contrasted this dynamic with face-to-face interactions: “*We know [the programme facilitator], so if [they] asked ‘What do you think?’, I would go ‘Well, I wasn’t too sure,’ whereas you might actually want to say you really didn’t like that bit… you don’t want to hurt anybody’s feelings.’ (PPT9)*. In this way, the quasi-human quality inherent in the chat-agent allowed some users to overcome a bias for social desirability to provide data that was more authentic to their true experience.

## 4 Discussion

### Summary of findings

In terms of feasibility (Aim A), CLEA operated with high fidelity across the 12-week deployment; consistently generating clear, relevant questions aligned to instructed scope. Completions of data collection sessions were high (80%), without substantial decline over time. One third of data collection sessions were deferred by 12 hours or more.

With respect to acceptability and usability (Aim B), the WhatsApp interface was experienced as familiar, accessible, and frictionless, lowering barriers to participation. Participants valued the reflective manner and affiliative tone of the chat-agent; expressing positive sentiment and reporting a sense of rapport promoting engagement longitudinally, despite requiring an investment of cognitive effort. While participants varied in their tendency to anthropomorphise CLEA, it was only when moments of robotic interaction occurred that engagement was reportedly undermined.

Regarding data quality and utility (Aim C), participants typically provided brief, highly relevant responses, typically focused on descriptions of recent experiences or events. While individual responses were limited in cognitive or emotional depth, useful insights were consistently generated at the session-level across the study period. Participants reported that personally relevant questions and a sense of non-judgement led to more open disclosure, affecting data quality.

In addition, participants appeared to co-construct accounts with the chat-agent, meaning data could be influenced by the systems’ conversational ability. Interventional effects were also observed (e.g, social support), influencing the research phenomenon under investigation, while reportedly promoting engagement.

### Implications of design choices

A keystone design choice within this implementation of CLEA was selecting the LLM powering its ability to generate text and ask questions. Model choice has *practical* implications in terms of cost and ease of technical integration. There are also *performance* implications; models vary in their ability to abide instruction, maintain conversational coherence, accurately interpret participant input and respond in a timely manner [36, 37, 38]. OpenAI’s GPT4-Turbo model was selected at the time as the optimal choice given these considerations. However, the significance of the model’s positive, reinforcing and encouraging interactional manner was unprecedented in influencing engagement and shaping data. Specifically, this surfaced through several cross-cutting interactional effects: the potential incitement of positive or elaborated responses [39]; ‘coaching-like’ interventional effects with potential to shape the phenomenon of interest over time; and the establishment and sustenance of rapport in promoting engagement and supporting disclosure. We noted that these effects create tension: the affiliative quality enhances engagement – through providing a sense of social support - but diminishes neutrality, thereby affecting data quality. Model choice is therefore a methodological decision with downstream implications for interaction, engagement, and data production.

Beyond underlying model tendencies, the content and style of chat-agent generated messages were largely governed by the system-level instructions (see 2.4) provided through prompt engineering. First, the chat-agent was instructed to adopt the role of a ‘reflective interviewer’. This choice reflected our pragmatic orientation and use of an interpretive lens for data collection in this context. Second, specifying the research scope within the system instructions establishes the goals that inform how the chat-agent directs inquiry. As such, careful attention in phrasing is required to ensure alignment and avoid embedding assumptions. Third, explicit instructions were implemented to restrict conversational turns (5 questions per session) and to only ask 2 follow-up questions on a specific topic within a session, before moving on. While primarily intended to manage burden and reduce repetition and fatigue, these constraints offer a partial explanation for the limited depth observed in Section 3.4.2; although, consistently eliciting reflective depth within a small number of questions is challenging even for experienced interviewers [40]. These conversational constraints therefore represent a deliberate methodological balancing act between participant burden, topical coverage, and depth of content.

The use of instant messaging in the form of WhatsApp was another design choice that can influence the nature of responses in particular ways. WhatsApp was selected for its familiarity, existing widespread adoption, and ease of use simplifying onboarding and ongoing use [41] . As is typical of everyday messaging communication practices [42], we also observed how typed responses to CLEA were mostly clipped and short. The limited reflective depth of participants’ responses is therefore unsurprising; it is unusual to convey substantial cognitive and emotional depth in a single WhatsApp reply. However, when considered at the level of the data collection session rather than individual replies, the analytic yield presents a different pattern. Nearly every session generated useful data in aggregate, across replies, producing new insights relevant to the research questions (Fig 6). Therefore, ‘complete’ responses appeared to emerge over message sequences. This is potentially explained by the phenomenon of externalising thinking to the co-constructive nature of quick successive interactional turns with a conversational partner (here, AI) in instant-messaging [42]. Additionally, WhatsApp users can have different practices for replying – sometimes responding ‘in the moment’, while other times delaying response. Interface choice therefore has connotations for the structure, content and momentariness of data.

### Guidance for reflexive design practice

CLEA is a high-level longitudinal research method that can be adapted through specific design choices to conduct ILR longitudinal research with varying qualities of EMA or diary method. Reflexive consideration of these design choices must be made, aligned to the requirements of the respective study context. In the table below we build on the concept of technological reflexivity, introduced by Paulus et al. [43] and applied to AI-assisted qualitative analysis by Prahl et al. [44]. We respond by operationalising its application to AI-assisted data collection. Using our findings, we identify the key design areas that warrant technological reflexivity in future CLEA adaptations. We emphasise there are trade-offs in design choice for researchers’ consideration. Table 2 presents key relevant design parameters and the implications these have on data.

**Table 2.** CLEA Design Implications for technological reflexivity.

| Design parameter | Methodological considerations and trade-offs for qualitative intensive longitudinal research | Future work |
| --- | --- | --- |
| Instant messaging interface | Existing mobile instant messaging (MIM) platforms (e.g., WhatsApp, Messenger) offer high familiarity, minimal onboarding burden, and proven usability, supporting accessibility and sustained engagement. However, established platform conventions (e.g., brevity of replies, informal tone) shape interactional norms with implications for the form, content, and immediacy of responses.<br>Custom-built interfaces afford greater control over interactional norms and data structure but increase onboarding friction and risk reduced engagement. | How data elicitation through CLEA varies by interface type (existing vs custom)<br>How interaction modalities (e.g., text vs voice input) affect disclosure, reflection, and response depth.<br>How response-window policies and notification practices interact with platform norms to shape temporal sensitivity and missingness. |
| AI model choice | Tone and interactional tendencies of the underlying AI model: affiliative and encouraging styles may foster rapport, reduce perceived judgement, and support longitudinal engagement. However, such positivity can introduce non-neutral feedback, potentially inciting inauthentic positivity, reshaping self-perception over time, or producing interventional effects through relational support. More neutral models may preserve analytic distance, but risk reduced acceptability and engagement.<br>Additional performance factors—including instruction-following, conversational coherence, latency, and safety behaviour—also affect data quality and interactional flow. | Compare CLEA deployments using LLMs that vary in affiliative versus neutral interactional styles while holding prompts constant, examining how differences in model manner influence engagement, disclosure, and longitudinal narrative construction, and explicitly report model versioning and stability as part of methodological transparency. |
| Prompt engineering: role designation | • Explicitly defining the conversational role of the chat-agent (e.g., reflective interviewer) embeds an epistemic stance into the data collection process. Instructing CLEA to operate through an interpretivist or constructivist lens may enhance meaning-making and reflective engagement but increases the likelihood of co-construction and reactivity.<br>• More neutral role designations may support standardisation but limit interpretive richness. Researchers' epistemological commitments therefore extend to system-level instructions governing CLEA's interactional behaviour. | • Evaluate how CLEA can be aligned with different research paradigms by systematically varying role designation (e.g., interpretivist, phenomenological, descriptivist) and examining downstream effects on interaction, engagement, and data characteristics.<br>• Reflexive reporting of epistemic assumptions embedded in system prompts should become standard practice. |
| Prompt engineering: scope of inquiry | • Defining the scope of inquiry (e.g., event-contingent experiences versus continuously fluctuating states) shapes which constructs can be meaningfully captured longitudinally.<br>• It can also embed assumptions about the participant or their experiences. Careful scope phrasing is therefore required to avoid pre-structuring or constraining participants' accounts. | • Examine how CLEA performs under different scope formulations targeting distinct phenomena (e.g., discrete events versus ongoing behavioural states).<br>• One promising direction is combining low-burden EMA-style inputs with contingent conversational reflection to integrate temporal sensitivity with interpretative sense-making. |
| Prompt engineering: interactional guidance | • Conversational constraints—such as limits on questions per session, follow-ups per topic, and topic switching—represent a balancing act between participant burden, breadth of coverage, and depth of elicitation.<br>• Restrictive probing can reduce fatigue and support sustained engagement but may constrain opportunities to explore ambiguity, contradiction, or emergent themes within sessions. | • Experiment with varying conversational constraints (e.g., number of follow-ups, adaptive probing) to identify configurations that optimise analytic yield, while preserving manageable participant burden. |

### How conversations unfolded: conversational dynamics, adaptive probing, and participant attitudes towards AI

Participants’ engagement with these conversations also depended on cognitive readiness. Many described needing sufficient “headspace” and an appropriate setting to respond thoughtfully, which sometimes led to deferred responses. This has implications for sampling - with deferral of responses until moments perceived as suitable, participants introduce self-selection into the timing of data provision. Deferred or delayed responses are also pervasive in both diary and EMA studies [45, 46], but can be mitigated in EMA designs that rely on random prompting with cut-offs for response windows. As a result, CLEA data may disproportionately represent contexts conducive to reflection, and missingness may not be random. Importantly, however, the cognitive investment was not framed as burdensome by participants but as a condition for engaging well. Over time, participants reported deriving value from the opportunity to reflect on personally meaningful aspects of their experience, which appeared to support sustained longitudinal engagement.

### Reactivity, interventional effects and other methodological limitations

CLEA is subject to the same forms of reactivity documented in EMA and diary research. Repeated assessment can increase self-awareness and influence participants’ thoughts, emotions, or behaviours through the very act of reporting [10, 14, 47]. In this respect, CLEA shares this effect with other intensive longitudinal methods; an important limitation for research through a positivist lens, in terms of removing perceived sources of bias. Standard EMA approaches often seek to reduce such reactivity through brief, neutral, and minimally intrusive prompts [10].

However, such strategies would be at odds with CLEA’s ability to promote longitudinal engagement through inviting participants to pause, reflect, and articulate experiences in language, making some degree of reactivity unavoidable. This could explain why CLEA did not show the longitudinal drop off characteristic of typical EMA studies [13]. In addition, CLEA’s conversational structure limits the feasibility of high-frequency sampling. EMA protocols often use very brief, low-effort prompts — including “micro-EMA” designs — to capture rapidly fluctuating states [10]. The cognitive effort involved in CLEA makes it less suited to such dense sampling strategies. CLEA is therefore better aligned with lower-frequency designs that prioritise interpretive depth over momentary closeness.

The longitudinal compound effect of reactivity may have augmented the health programme itself over time; participants noted, for example, interacting with CLEA helped consolidate learnings from programme sessions. Additionally, accounts suggest that the repeated act of articulating experiences in response to CLEA prompts could encourage processes of sense-making and heighten awareness of progress and challenges. These effects align with well-established mechanisms in self-monitoring and reflective practice, whereby attention and articulation can themselves contribute to behavioural change [6, 14]. In this respect, CLEA may participate in shaping the phenomenon under study, through cognitive processes associated with repeated reflection.

### Positioning CLEA relative to other intensive longitudinal research methods

Our findings position CLEA as methodologically distinct from EMA and diary and interview methods. CLEA appears Like EMA, CLEA can be configured to capture experiences close to their occurrence and anchored in context. However, typical EMA designs could be better suited for exploring highly temporally sensitive phenomena such as pain or cravings, or studies looking to model dynamics which can change over minutes or hours [48]. Like diary methods, CLEA relies on reflective engagement and meaning-making over time [6, 8]. CLEA extends this through its conversational and adaptive format, enabling inquiry to unfold dynamically in response to participant input rather than being fully specified in advance. While CLEA can gather relatively short responses describing specific and temporally sensitive insights, more traditional diary methods—which afford participants greater freedom to determine the scope and structure of entries—may be better suited to long-form narrative and broad, participant-led accounts of experience [7]. In comparison, our instance of CLEA appears better aligned to longitudinal sense-making of specific phenomenon somewhat narrower in scope; where shorter bursts of data capture lower participant burden, improve compliance, and yield more focused data for analysis. Similarly, considering the content of data CLEA elicits, interviews are likely most appropriate for studies seeking to understand emotionally complex issues. However, to address issues of recall in the retrospective interview, a hybrid approach utilising CLEA for gathering specific insights longitudinally could be used to prompt recollection, as per Kwasnicka et al. [51].

### Limitations and future work

Several limitations should be considered when interpreting these findings. First, we sampled a relatively small and demographically homogeneous group. Future work is required to understand the generalisability of the findings and how the technology would need to be adapted for other populations, for example paediatric populations, where EMA protocols need to be adapted to meet the distinct developmental characteristics [49]. Second, we operationalised only a single configuration of CLEA. Work is needed to examine the effects of reflexive adjustment of key design parameters, including model selection, prompt engineering, and user interface choice (see Table 2). Third, measures of response depth were necessarily approximate and used to provide some insight into the type of data elicited. While we used a specific, literature informed measure to approximate reflective depth, such an approach cannot fully characterise the concept of ‘depth’ in qualitative research or convey the value conversational data. Fourth, participants entered the study at slightly different time points, resulting in variation in exposure length.

Although this reflects real-world implementation conditions, it may have resulted in uneven data contributions across participants. Finally, CLEA was embedded within an ongoing health programme, and engagement in the programme itself may have influenced participants’ responsiveness and motivation to engage with CLEA. As such, future work should investigate if engagement varies when deployed standalone.

## 5 Conclusion

These findings introduce CLEA as a viable, ecological method for longitudinal qualitative health research using conversational AI. CLEA was acceptable and engaging for a lower socioeconomic group involved in a weightloss programme. It was capable of capturing useful insights into their programme experience and associated behaviour change, over time. CLEA offers both benefits and trade-offs for researchers considering longitudinal research methods. Its accessible, highly engaging user experience is well suited to longitudinal protocols, though findings on limited reflective depth indicate it is not a substitute for the emotional depth achievable through skilled human interviewing. Compared with typical EMA designs, CLEA may enable greater interpretative richness through conversational interaction, but with reduced momentariness. Diary methods likely offer more open-ended, self-led reflection, but often at the cost of reduced topical relevance and increased analytic burden. Though, comparative studies are required to substantiate these comparisons. Rather than replacing existing approaches, CLEA may best be understood as complementary to them. Hybrid EMA–CLEA designs may enable interpretive reflection anchored in momentary data, while CLEA-derived longitudinal insights could support recall and sense-making in retrospective interviews. Importantly, this study highlights the particularly consequential design choices that affect acceptability and engagement with CLEA, aswell as the form and content of data it produces. We therefore urge reflexivity in the development and reporting of future CLEA implementations, attending carefully to technological design decisions. We hope this work provides practical guidance for extending CLEA with caution, rigour, and transparency.

## Data Availability

Considering the highly sensitive personal qualitative data in our study, in line with participant data protection due to risk of re-identification, our research data is not available on a publicly held data repository. However, anonymised data is available on request, held in the university of Bristol's research data storage facility https://www.bristol.ac.uk/acrc/research-data-storage-facility/ Requests for partial or completely anonymised data may be made to the corresponding author

## Acknowledgements

We are grateful to the Bristol City Robins Foundation for their thoughtful guidance in designing accessible technology and facilitating warm introductions to participants.

Our research participants welcomed us into their group and beyond their active participation helped make this an enjoyable project, for which we give thanks.

